# Sex-specific differences in physiological parameters related to SARS-CoV-2 infections among a national cohort (COVI-GAPP study)

**DOI:** 10.1101/2023.09.17.23295693

**Authors:** Kirsten Grossmann, Martin Risch, Andjela Markovic, Stefanie Aeschbacher, Ornella C Weideli, Laura Velez, Marc Kovac, Fiona Pereira, Nadia Wohlwend, Corina Risch, Dorothea Hillmann, Thomas Lung, Harald Renz, Raphael Twerenbold, Martina Rothenbühler, Daniel Leibovitz, Vladimir Kovacevic, Paul Klaver, Timo B Brakenhoff, Billy Franks, Marianna Mitratza, George S Downward, Ariel Dowling, Santiago Montes, Duco Veen, Diederick E Grobbee, Maureen Cronin, David Conen, Brianna M Goodale, Lorenz Risch, the COVID-19 remote early detection (COVID-RED) consortium

## Abstract

Considering sex as a biological variable in modern digital health solutions, we investigated sex-specific differences in the trajectory of four physiological parameters across a COVID-19 infection.

A wearable medical device measured breathing rate, heart rate, heart rate variability, and wrist skin temperature in 1163 participants (mean age = 44.1 years, standard deviation [SD]=5.6; 667 [57%] females). Participants reported daily symptoms and confounders in a complementary app. A machine learning algorithm retrospectively ingested daily biophysical parameters to detect COVID-19 infections. COVID-19 serology samples were collected from all participants at baseline and follow-up. We analysed potential sex-specific differences in physiology and antibody titres using multilevel modelling and t-tests.

Over 1.5 million hours of physiological data were recorded. During the symptomatic period of infection, men demonstrated larger increases in skin temperature, breathing rate and heart rate as well as larger decreases in heart rate variability than women. The COVID-19 infection detection algorithm performed similarly well for men and women.

Our study belongs to the first research to provide evidence for differential physiological responses to COVID-19 between females and males, highlighting the potential of wearable technology to inform future precision medicine approaches.

This work has received support from the Princely House of the Principality of Liechtenstein, the government of the Principality of Liechtenstein, the Hanela Foundation in Switzerland, and the Innovative Medicines Initiative (IMI) 2 Joint Undertaking under grant agreement No 101005177. This Joint Undertaking receives support from the European Union’s Horizon 2020 research and innovation programme and EFPIA.

## Introduction

On March 11, 2020, the WHO declared the fast spreading coronavirus disease (COVID-19) a global pandemic [1]. This novel viral disease was first detected in Wuhan, China in December 2019 and is caused by severe acute respiratory syndrome coronavirus (SARS-CoV-2) [2]. Increasing knowledge about risk factors and symptoms, as well as the implementation of mass reverse transcription polymerase chain reaction (RT-PCR), serological tests, vaccines and social restrictions have helped control its spread [3,4]. However, asymptomatic virus transmissions and emerging virus mutations pose ongoing challenges in dealing with the pandemic. Today, more than two years after the first case was detected, many countries worldwide continue to experience waves of rising infections, with numerous unknowns remaining in our understanding of SARS-CoV-2. In particular, consistent data about the role of sex in relation to COVID-19 are lacking [5,6]. Significant changes in physiological parameters as breathing rate, heart rate, heart rate variability, and wrist skin temperature during a COVID-19 infection [7] raise the question about sex-specific differences within the trajectory of these parameters. A better understanding of sex-specific trajectories in physiological responses to the infection may support early detection and treatment of COVID-19.

A meta-analysis found that men with COVID-19 were globally almost three times more likely than women to be admitted to an intensive treatment unit [8]. Furthermore, the disease’s mortality rates were higher in men [9], potentially due to sex-specific differences in angiotensin converting enzyme 2 (ACE2) expression [10,11]. On the other hand, women were found to more frequently experience persistent symptoms such as dyspnoea and fatigue several months after the acute phase of the illness [12]. The infection rates were similar between the sexes [8], although this observation may differ between countries [13]. Moreover, initial analyses of eumenorrheic women’s susceptibility to SARS-CoV-2 among a real-world sample are in line with previously shown immune function fluctuations across the menstrual cycle [14] and suggest increased susceptibility during the luteal phase [15]. Research on sex-specific differences in immune responses that underlie COVID-19 disease outcomes showed higher plasma levels of innate immune cytokines such as IL-8 and IL-18 along with more robust induction of non-classical monocytes in male patients, whereas female patients showed higher T cell activation during SARS-CoV-2 infection [16]. Also, higher levels of innate immune cytokines were associated with worse disease progression in female patients [16].

To the best of our knowledge, our work represents the first investigation of sex-specific differences in SARS-CoV-2 affected physiological parameters as measured by a medical device. Previous studies have shown that direct-to-consumer and easy to use products with wide market availability such as Fitbit [17], smartwatches [18], the Ava bracelet [7,19], and other wearable devices [20] could be used for surveillance of changes in physiological parameters to give the user an early warning before COVID-19 symptom occurrence [21] or during asymptomatic infection [22]. The COVI-GAPP study investigated the applicability of the Ava bracelet for pre-symptomatic detection of COVID-19 [23]. Developed as a fertility tracker, the bracelet measures physiological parameters including wrist skin temperature, breathing rate, heart rate, heart rate variability and skin perfusion [24]. The previously published interim analysis of the COVI-GAPP dataset demonstrated significant changes in skin temperature, breathing rate, heart rate and heart rate variability during a COVID-19 infection [7]. These parameters were used to develop a machine learning (ML) algorithm for detection of pre-symptomatic SARS-CoV-2 infection which successfully detected 68% of COVID-19 cases up to two days before symptom onset. The algorithm is currently being tested and validated in a larger population with real-time access to the algorithm’s predictions [19].

The current work analyzed the same physiological parameters collected in the COVI-GAPP study to quantify sex-specific differences before, during and after a COVID-19 infection. We examined differences in trajectories of physiological parameters over five defined phases (baseline, incubation, pre-symptomatic, symptomatic, recovery) between female and male participants. Furthermore, we evaluated the performance of our ML algorithm for female and male participants separately with the goal to assess and correct a potential sex bias in its functionality.

## Materials and Methods

### Study design and participants

Since 2010, the observational population-based Genetic and Phenotypic Determinants of Blood Pressure and Other Cardiovascular Risk Factors (GAPP) study aims to better understand the development of cardiovascular risk factors in the general population of healthy adults aged 25 to 41 years [25]. From 2170 GAPP participants, 1163 individuals were enrolled in the COVI-GAPP study with inclusion and exclusion criteria published previously [23]. Data were collected from April 14, 2020, until January 31, 2022. The local ethics committee (KEK, Zürich, Switzerland) approved the study protocol, and written informed consent was obtained from each participant prior to enrolment (BASEC 2020-00786).

### Data collection

Physiological parameters of interest for this analysis were breathing rate, heart rate, heart rate variability, and wrist skin temperature, measured every 10 seconds by a wrist-worn bracelet while the user slept. The CE-certified and FDA-cleared Ava Fertility Tracker (version 2.0; Ava AG, Switzerland) was originally built to detect ovulating women’s fertile days in real time with 90% accuracy [26–28]. The bracelet’s three sensors can track biophysical changes regardless of the wearer’s sex [7] and was used in this study for detecting infection-based deviations from baseline parameters in both men and women (regardless of their menstruating status). Participants synchronized their bracelet each morning upon waking to a complementary smartphone app.

In addition to automatically collected physiological data, participants also provided information in the complementary app about their daily alcohol, medication, and drug intake (Fig 1A), as these substances can alter central nervous system functioning [29]. Furthermore, the app included a customized user functionality where participants reported COVID-19 symptoms in a daily diary (Fig 1B). Participants were also able to see and monitor changes in their physiological parameters in the app.

**Fig 1.**
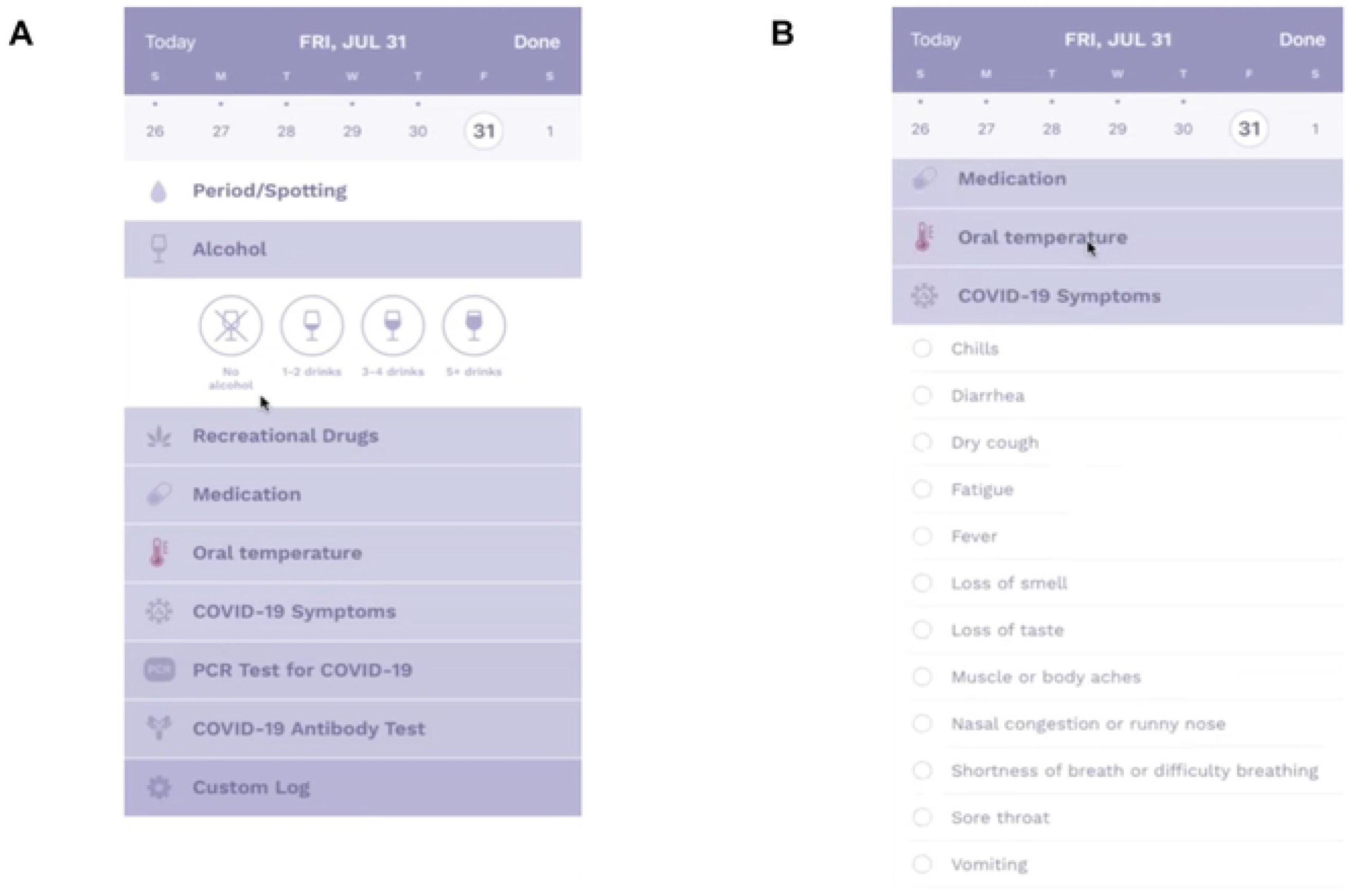
Participants wore a wrist-based medical device at night. Upon waking, participants synchronized the device with a complementary smartphone application and reported alcohol, medication, and drug intake (A) as well as potential COVID-19 symptoms (B) in the app.

### SARS-CoV-2 antibody testing and RT-PCR testing

SARS-CoV-2 antibody tests were performed by the medical laboratory Dr Risch Ostschweiz AG (Buchs SG, Switzerland) with an orthogonal test algorithm employing electrochemiluminescence (ECLIA) assays testing for pan-immunoglobulins directed against the N antigen and the receptor binding domain (RBD) of the SARS-CoV-2 spike protein, as described by Risch et al [30]. The enacted procedure ensures testing for actual SARS-CoV-2 infection independent of vaccine status. Baseline data were collected starting in April 2020 onwards (run 1; R1). Three follow-up blood samples (run 2, R2; run 3, R3; and run 4, R4) were collected within the scope of the study (Fig 2). The cut-off levels used for positive and negative values were ≥ 1.0 and ≤ 0.1, respectively. Values between 0.2 – 0.9 were considered as gray zone. Seroconversion was assumed if the first blood sample was negative for SARS-CoV-2 antibodies, but a subsequent sample was positive. Follow-up calls with participants who tested positive were performed to discuss their symptoms and duration.

**Fig 2.**
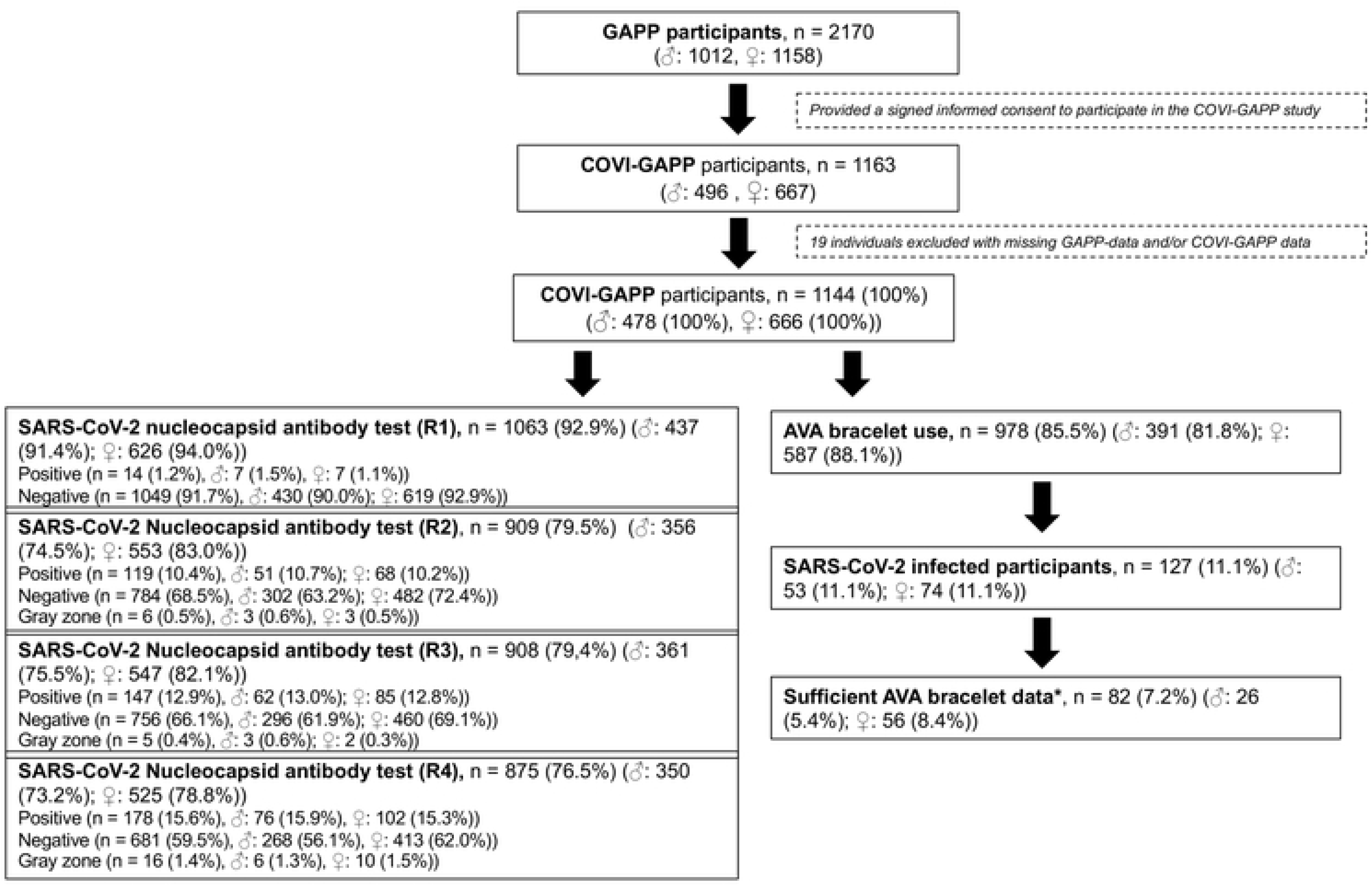
Study flow chart of the 1,163 participants that are enrolled in the COVI-GAPP study. The cut-off levels used for positive and negative values were ≥ 1.0 and ≤ 0.1, respectively. Values between 0.2 – 0.9 were considered as gray zone * Successful bracelet synchronization on more than 50% of days around symptom onset.

### Questionnaires

When visiting the study centre for SARS-CoV-2 antibody tests, participants were asked to answer a questionnaire about their personal information (age, sex), smoking status (current, past, never), as well as symptoms and hospitalizations during COVID-19 infection. Body mass index (BMI) based on height and weights were calculated with data from the GAPP database.

### Statistical Analysis

Our primary objective was to examine sex differences in the trajectory of breathing rate, heart rate, heart rate variability, and wrist skin temperature across a SARS-CoV-2 infection. Secondarily, we evaluated a machine learning algorithm designed for early detection of COVID-19 separately in male and female participants to examine potential sex biases in algorithm performance. Furthermore, we assessed sex-specific differences in antibody titres after SARS-CoV-2 infections. We processed and analysed all data using R (version 4.1.1) [31] and Python (version 3.6) [32].

#### 1. Sex-specific differences in COVID-19 related physiological parameters

To examine the association between sex and physiological parameters during baseline, incubation, pre-symptomatic, symptomatic and recovery phases of a COVID-19 infection, we applied multilevel linear mixed models with random intercepts and slopes including residual maximum likelihood estimation (REML) and Satterthwaite degrees of freedom. A multiplicative interaction term tested the association between sex and infection phase. All signals measured more than 10 days before symptom onset via phone call confirmation with a study team member were categorized as occurring during the baseline period. The incubation period was defined as the time interval from 10 days up to 3 days before symptom onset. The pre-symptomatic period was defined as the two days before symptom onset, while the symptomatic period lasted from the day of symptom onset until the day symptoms ended. All signals measured after symptom end were categorized as occurring during the recovery period. We dummy coded four variables to indicate the period within which the signal occurred, with baseline serving as the reference period. Each of the four multilevel models was compared to the corresponding null model (i.e., an intercept-only model) by means of an ANOVA.

As a sensitivity analysis, we also tested potentially confounding variables as single terms in additional models to determine whether changes in physiological parameters occurred due to COVID-19 infection over and above changes associated with participant age, BMI, hypertension, medication, alcohol, and recreational drugs.

#### 2. Sex-specific differences in algorithm’s performance

The retrospective ML algorithm, developed as described in previous papers, [7,19] aimed to detect a COVID-19 infection prior to symptom onset. The algorithm was designed to ingest trends in physiological signals across sets of days to detect deviations in these signals and predict a potential infection. The model was trained to predict infection two days and one day prior to symptom onset, as well as on the day of symptom onset. Here, we assessed the algorithm’s performance metrics separately for male and female COVI-GAPP participants to identify any potential sex bias in the model. Performance metrics were calculated per day in participants who tested positive where days from -40 to -2 relative to the onset of the first symptoms were considered negative and days -2 to day 0 as positive. In other words, positive predictions of the algorithm prior to 2 days before symptom onset, these predictions were interpreted as false positives. The set of metrics selected for the evaluation of the algorithm included precision (number of true positives divided by sum of true positives and false positives), recall (number of true positives divided by the sum of true positives and false negatives), and F-score (the harmonic mean of precision and recall).

#### 3. Sex-specific differences in antibody titres of SARS-CoV-2 Nucleocapsid after COVID-19 infection

To enable a reliable comparison of antibody titres after a COVID-19 infection, antibody titres (values > 1.0) against the SARS-CoV-2 Nucleocapsid were compared between sex. Blood was collected four times over the course of the study with varying sample sizes (Fig 2). Normally distributed continuous variables were compared using unpaired t-tests, and non-parametric continuous variables were compared using Mann-Whitney U tests.

## Results

### Participants

A total of 1163 participants (mean age = 44.1 years, standard deviation [SD] = 5.6; 667 [57%] females) were enrolled in the study. During the study period, 127 participants (10.9%; [9.3,12.8]) contracted COVID-19. Eighty-two participants (mean age = 42.6 ± 5.3 years; 56 [68%] females) testing positive for SARS-CoV-2 had worn and synchronized their bracelet successfully on more than 50% of days around symptom onset (i.e., at least 20 days before and 20 days after symptom onset), thereby ensuring sufficient quality of data to be included in analyses. The number of days with successfully synchronized bracelet data did not differ (*p* = 0.967) between females (range 67 to 511 days; mean = 239.6 ± 71.8 days) and males (range 45 to 508 days; mean = 238.8 ± 86.4 days). With regards to the reported symptom duration, values for four participants (2 females) were missing and imputed based on the median across the sample.

Blood samples and questionnaire data were available from 1,144 participants. Mean age and BMI of these participants were 45 (± 5.5) and 24.7 (± 3.9), respectively. At baseline, male participants had significantly higher BMIs (26.17 ± 3.41) than female participants (23.70 ± 3.96; t(1079) = 10.71, *p*<0.001). They also reported significantly higher rates of hypertension (7.74%) than female participants (3.15%; *X*^2^(1) = 11.23, *p<*0.001). Analyses did not reveal any significant sex-based differences in smoking status, age, or hospitalization rate (Table 1).

**Table 1.** Baseline characteristics stratified according to sex were collected by questionnaires completed within the GAPP study. Information about hospitalization were collected 4 times (01 – 04) as part of a questionnaire for SARS-CoV-2 positive participants within the COVI-GAPP study. Data are presented as mean ± SD, or number.

| Variables | Total<br>n = 1,144 | Male<br>n = 478 | Female<br>n = 666 | Test statistics | Significance<br>(p value) |
| --- | --- | --- | --- | --- | --- |
| Smoking status, N<br>(never: current: past<br>smoker) | 658: 167:<br>319 | 265: 68:<br>145 | 393: 99:<br>174 | X2 (2) = 2.46 | 0.292 |
| Hypertension, N (yes: no) | 58: 1086 | 37: 441 | 21: 645 | X2 (1) = 11.23 | <0.001 |
| Age, years (±SD) | 43.99 (±<br>5.51) | 44.3<br>(±5.35) | 43.77<br>(±5.61) | t (1057) =<br>1.53 | 0.1449 |
| BMI, kg/m2 (±SD) | 24.72<br>(±3.94) | 26.17<br>(±3.41) | 23.7<br>(±3.96) | t (1079) =<br>10.71 | <0.001 |
| Hospitalization 01, N (yes:<br>no) | 0:10 | 0:4 | 0:6 | Fisher's exact<br>test | 1 |
| Hospitalization 02, N (yes:<br>no) | 11:113 | 7:44 | 4:69 | X2 (1) = 2.52 | 0.2047 |
| Hospitalization 03, N (yes:<br>no) | 2:23 | 0:12 | 2:11 | X2 (1) = 0.46 | 0.4973 |
| Hospitalization 04, N (yes:<br>no) | 3:47 | 0:24 | 3:23 | X2 (1) = 1.25 | 0.2625 |

#### Sex-specific differences in COVID-19 related physiological parameters

We show the trajectory each of the four analysed physiological parameters during a SARS-CoV-2 infection separated by sex (Fig 3). The multilevel models revealed significant differences between male and female participants in all parameters during the symptomatic period (Table 2). We observed a larger increase in skin temperature, breathing rate and heart rate as well as a larger decrease in heart rate variability in males compared to females during this period. Moreover, male participants’ breathing rate and heart rate remained at significantly higher levels during the recovery period as compared to their female peers (Table 2). Each of the four models provided a significantly better fit to the data than the corresponding null model (p<0.0001).

**Fig 3.**
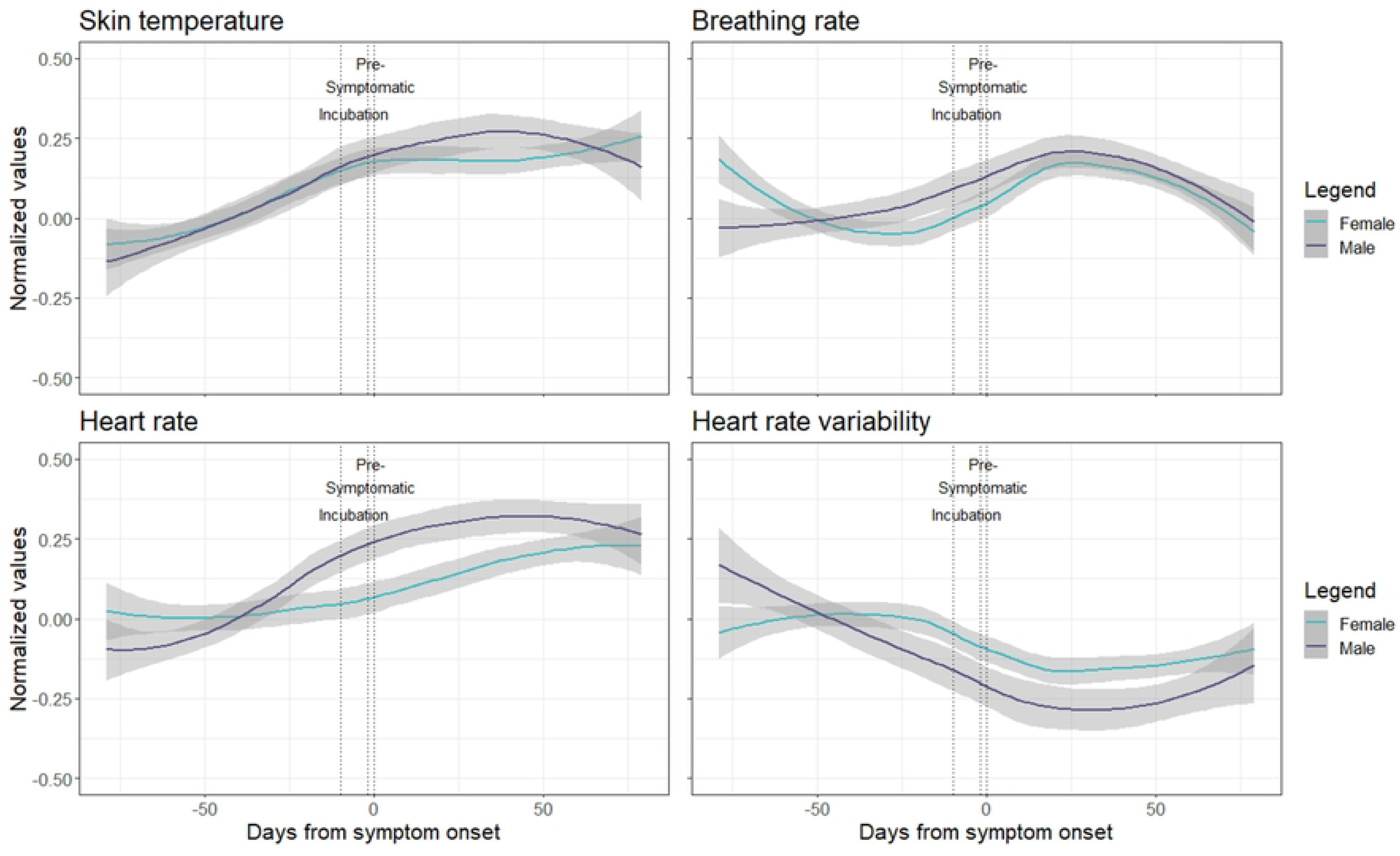
Trajectory of the four analysed physiological parameters across the course of a confirmed COVID-19 infection, centred around participant-reported symptom onset. The values of each physiological parameter (with 95% CIs) were normalized according to each individual’s baseline measurements and collapsed across females (n=56) and males (n=26).

**Table 2.** Results from multilevel linear mixed models showing the main effects of infection phase and sex as well as the interactions between the two with regards to changes in physiological signals. Unstandardized beta coefficients are presented, with *p*-values in parentheses and in bold if lower than 0.05. Sex was coded such that positive coefficients represent larger values in females.

|  | Skin temperature | Breathing rate | Heart rate | Heart rate variability |
| --- | --- | --- | --- | --- |
| Intercept | 35.01 (<0.0001) | 13.51 (<0.0001) | 46.98 (<0.0001) | 4.3 (<0.0001) |
| Infection phase |  |  |  |  |
| Baseline | Reference | Reference | Reference | Reference |
| Incubation | 0.18 (0.15) | 0.33 (0.13) | 1.49 (0.12) | -0.25 (0.12) |
| Pre-symptomatic | 0.23 (0.26) | 0.71 (0.17) | 1.26 (0.41) | -0.18 (0.42) |
| Symptomatic | 0.74 (<0.0001) | 2.93 (<0.0001) | 6.88 (<0.0001) | -0.93 (<0.0001) |
| Recovery | 0.22 (0.0006) | 0.38 (0.004) | 2.17 (0.003) | -0.28 (0.09) |
| Sex, female | 0.45 (<0.0001) | 0.91 (0.06) | 4.96 (0.001) | -1.35 (<0.0001) |
| Interaction |  |  |  |  |
| Sex*Incubation | -0.02 (0.74) | -0.2 (0.11) | -0.36 (0.5) | 0.09 (0.34) |
| Sex*Pre-symptomatic | -0.01 (0.92) | -0.26 (0.38) | 0.07 (0.93) | 0.04 (0.78) |
| Sex*Symptomatic | -0.28 (<0.0001) | -1.31 (<0.0001) | -3.09 (0.0001) | 0.43 (<0.0001) |
| Sex*Recovery | -0.04 (0.23) | -0.25 (0.001) | -0.96 (0.02) | 0.11 (0.25) |

When including age, BMI, hypertension diagnosis, medication, alcohol and drug intake, the interactions between sex and phase of infection remained unchanged indicating that they cannot be explained by the influence of these confounding variables (S1 Table).

### Sex-specific differences in algorithm’s performance

Table 3 provides a by-sex breakdown for the algorithm’s performance. Sensitivity score can be found as the recall of the positive class (days with an existent SARS-CoV2 infection), while specificity is the recall of the negative class (days without a SARS-CoV2 infection). The algorithm showed the same precision (i.e., 92) when giving a SARS-CoV2 positive alert, across participant sex. Cross-class recall was more balanced among females than males in our sample. Detecting 53% of SARS-CoV-2 positive days in females, the algorithm performed less well in males (26% of SARS-CoV2 positive cases detected).

**Table 3.**
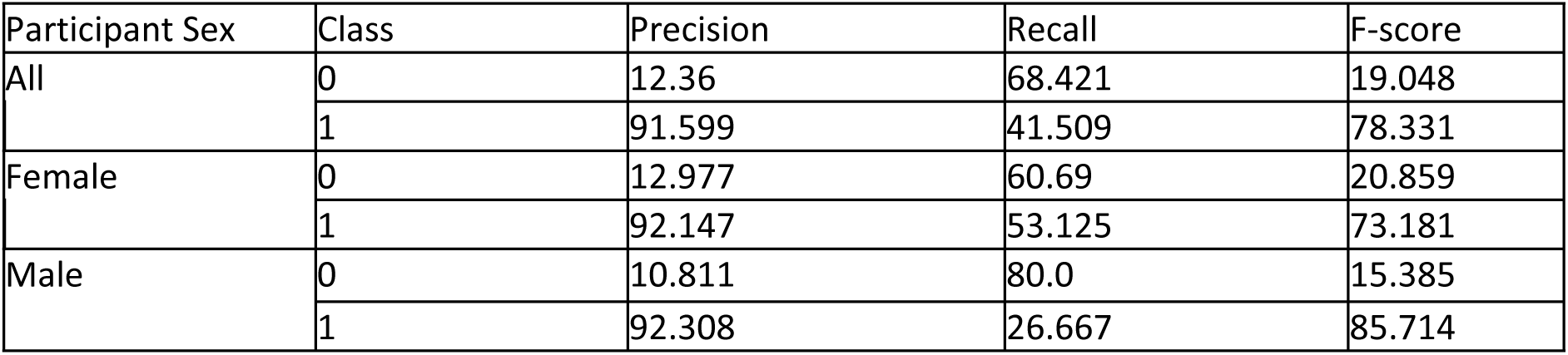
Performance metrics of the machine learning algorithm for female and male participants. Sensitivity score can be found as the recall of the positive class (i.e., days with an existent SARS-CoV2 infection), while specificity is the recall of the negative class (i.e., days without a SARS-CoV2 infection).

| Participant Sex | Class | Precision | Recall | F-score |
| --- | --- | --- | --- | --- |
| All | 0 | 12.36 | 68.421 | 19.048 |
|  | 1 | 91.599 | 41.509 | 78.331 |
| Female | 0 | 12.977 | 60.69 | 20.859 |
|  | 1 | 92.147 | 53.125 | 73.181 |
| Male | 0 | 10.811 | 80.0 | 15.385 |
|  | 1 | 92.308 | 26.667 | 85.714 |

### Sex-specific differences in antibody titres of SARS-CoV-2 Nucleocapsid after COVID-19 infection

Antibody titres of the female and male sub-groups were not significantly different across runs. Nucleocapsid antibody values in run 1 trended higher in male participants (Table 4).

**Table 4.** SARS-CoV-2 Nucleocapsid (N) antibody (AB) values stratified according to sex. data are presented as mean ± SD, or median (interquartile range).

**This is the Table 4 Title. SARS-CoV-2 Nucleocapsid (N) antibody (AB) values stratified according to sex.**
| Variables | Male (n = 7) | Female (n = 7) | Test statistics | Significance (p value) |
| --- | --- | --- | --- | --- |
| SARS-CoV-2 N AB run1 | 32.66 (± 34.74)a | 75.66 (± 46.87)a | W = 13 | 0.14 |
|  | Male (n = 51) | Female (n = 68) |  |  |
| SARS-CoV-2 N AB run2 | 34.1 (1 – 183.7)b | 33.4 (1.7 – 212.2)b | W = 1753 | 0.92 |
|  | Male (n = 62) | Female (n = 85) |  |  |
| SARS-CoV-2 N AB run3 | 40.05 (1 - 274) | 29.7 (1.4 – 234.3) | W = 2772 | 0.59 |
|  | Male (n = 76) | Female (n = 102) |  |  |
| SARS-CoV-2 N AB run4 | 17.95 (1.2 - 221) | 36.59 (1 – 266.4) | W = 4280 | 0.235 |

## Discussion

The presented study examined sex-specific differences in physiological parameters among 82 individuals with a documented SARS-CoV-2 infection. We found that male participants experienced significantly larger increases in wrist skin temperature, breathing rate and heart rate as well as larger decreases in heart rate variability during the symptomatic period compared to females. In one of the first prospective cohort studies relying on wearable sensor technology to collect real-time continuous physiological signals, we provide evidence for sex-based differential physiological responses to COVID- 19.

Considering the higher mortality and hospitalization rates observed in male COVID-19 patients [9], our findings may reflect sex-specific biological responses to the infection. In line with previous work [16], we did not observe any differences between the sexes with regards to antibody titers. However, Takahashi et al. [16] observed a stronger acute T-cell response in female as compared to male COVID- 19 patients. The poorer T-cell response in men was associated with their worse disease progression. On the other hand, the authors measured higher levels of several pro-inflammatory innate immunity chemokines and cytokines in men as compared to women. They thus concluded that the early phase of COVID-19 is associated with key sex differences in immunological mechanisms potentially accounting for the differential disease progression between women and men.

Given that the sex differences in physiological signals in our study are most pronounced during the symptomatic phase, we propose that they reflect the above mentioned sex-specific immunological mechanisms [33]. Inflammatory markers (e.g., cytokines) have been shown to reflect disease severity in COVID-19 [34]. As the autonomic nervous system is known to modulate inflammation [35] and the examined physiological signals reflect the function of the autonomic nervous system [36], our findings suggest support for differential immunological responses to COVID-19 between the sexes.

Importantly, altered physiological signals such as decreased heart rate variability and increased skin temperature have been proposed as prognostic markers for several disorders including cardiovascular disease [37] as well as infectious diseases like COVID-19 [18,38–40]. Modern wearable technology represents a unique and powerful framework to collect continuous real-time physiological data. The predictive value of physiological signals combined with the reliable history of measurements provided by wearables opens up new avenues to inform clinical actions and support future precision medicine approaches incorporating a variety of individual factors into clinical decisions (reviewed in Mitratza et al. [41]).

An important step towards precision medicine can be made by considering sex differences in modern digital health solutions. Historically, women have been underrepresented in clinical trials leading to medical solutions focusing on men at the risk to women’s health [42]. Many diseases differ between female and male patients with regards to the prevalence, progression or response to treatment [43]. For example, minor stroke is more often missed in female than male [44] patients, possibly due to definitions in clinical diagnosis reflecting typical manifestations in males [43]. More recently, a sex bias has been recognized in modern ML solutions that are often developed and trained on male data and thus result in better performance in men [45]. Therefore, in the presented work, we examined sex differences in the performance of our ML algorithm for early detection of COVID-19. The algorithm reached a higher sensitivity for female participants. We postulate this difference may be due to the larger sample size in the female group. However, the algorithm’s precision was the same in both groups indicating that it is suitable for use in both men and women, as intended.

## Limitations

While our study belongs to the first research to consider sex-based differences in COVID-19 detection using digital health, future work could continue to build upon our findings by examining the casual mechanism underlying differences between SARS-Cov-2 infected men and women. In particular, inability to disentangle immunological versus menstrual-driven changes in physiological parameters among female participants limits our research’s generalizability. In menstruating women, a specific pattern has been recognized in the trajectory of physiological signals across the menstrual cycle. Goodale et al. have demonstrated increased skin temperature, heart rate and breathing rate in the postovulatory phase of the cycle, mirroring cycle-based shifts in sex hormones [26]. Sex differences in physiological signals measured in the current study may thus partly be due to hormonal impact. We cannot exclude such influence as we had limited information about female participants’ menstrual cycle or reproductive health (e.g., usage of hormonal birth control, menopausal status). Future researchers may wish to record participants’ menstrual status and measure hormone levels directly, to probe the relationship between sex hormones and physiological differences.

Nevertheless, we believe that menses-driven changes in physiology do not adequately explain the sex differences in our results, as the dynamics of the observed physiological signals are in line with previous reports regarding COVID-19 and include increased skin temperature, heart rate and breathing rate as well as decreased heart rate variability during infection [20]. Furthermore, the most pronounced sex differences in our study occurred during the symptomatic period, suggesting a disease-triggered disparity among males and females. Moreover, we do not expect that the distribution of menstrual cycle phases follows a specific pattern for our participants. We rather expect it to be random, and thus the hormonal effects to be cancelled out. Finally, 30% (n=17) of females in the sample were older than 45 years; peri- or post-menopausal, they were beyond reproductive age and thus unlikely to experience menses-modulating effects on their physiological parameters.

Another limitation important to note is the potential effect of recall bias on our findings. The COVID- 19 symptom onset date was determined based on the participants’ retrospective reports, and the classification of the relevant infection periods (i.e., incubation, pre-symptomatic and symptomatic period) was based on this date. Therefore, an unreliable report would be associated with an inaccurate definition of the infection periods leading to shifts in trajectories of physiological signals. Furthermore, in the effort to smooth the data in the model, the abrupt changes in physiological signals after infection generated gradual alterations in the estimated trajectory. The deviations from the baseline during the first and last days may be reflective of such model artifacts (Fig 3). Finally, it is important to note that we did not adjust any parameters from our statistical tests to account for multiple testing. Therefore, we acknowledge chances for type 1 error in our findings. Nevertheless, we believe that our research provides important initial insights to be confirmed in future investigations.

## Conclusion

For the first time, we show sex differences in physiological responses to COVID-19. Our results highlight the importance of taking sex into account in medical treatment and care of COVID-19 patients, as well as when validating infection detection algorithms in digital health. Moreover, we reveal the potential of continuous real-time physiological signals as a clinical tool to inform future precision medicine approaches.

## Data Availability

The data underlying the results presented in the study are available from (Private University of the Principality of Liechtenstein, Institute for Laboratory Medicine, 9495 Triesen).

## Acknowledgements

We thank the GAPP participants who enrolled in this study. Additionally, the authors thank the following for their contributions to the study: The local study team in Vaduz, FL, the different teams at the Dr Risch medical laboratories in Vaduz and Buchs, CH. We would also like to thank the Coobx AG in Balzers, FL, for the provision of 3D printed bracelet extensions for persons with large wrists. Addressing data protection issues, we acknowledge the substantial collaborative support of the Elleta AG as well as the national data protection agency in Liechtenstein. We thank the government of the Principality of Liechtenstein, the health ministers, and the Liechtenstein Office of Public Health for their support. Finally, our thanks are especially due to the Princely House of Liechtenstein, which gave decisive support that enabled the initiation of this project.

## Supporting information

**S1 Table. This is the S1 Table Title. Results from multilevel linear mixed models showing the main effects of infection phase, sex, age, medication, drug and alcohol intake, BMI and hypertension as well as interactions between sex and infection phase with regards to changes in physiological signals.**

This is the S1 Table legend. Unstandardized beta coefficients are presented, with p-values in parentheses and in bold if lower than 0.05. Sex was coded such that positive coefficients represent larger values in females. Hypertension was a binary variable representing diagnosed hypertension. In addition to significant effects of infection phase and sex described in the main text, we observed a significant main effect of medication and alcohol intake on physiological signals. Nevertheless, these effects did not alter any of the multilevel model results reported in the main text.

## CRediT Statement for Authorship

All authors critically reviewed and approved the final version of this manuscript and had final responsibility for the decision to submit for publication.

Conceptualization: MRi, HR, RT, PK, TB, BF, MM, GD, AD, SM, MC, DEG, DC and BMG, LR; Data curation: KG, SA, MRo., VK, AM, and BMG; Formal analysis: MRi, KG, NW, OCW, MRo, DL, VK, AM, and BMG; Funding acquisition: MRi, MC, DEG, DC and LR; Investigation: KG, SA, OCW and MM; Methodology: MRi, AM, VK, KG, PK, TB, BF, GD, SM, MC, DEG, DC, BMG, and LR; Project administration: KG, OCW, SM, MC and DEG; Resources: MRi, KG, SA, OCW, MK, NW, CR, DH and TL; Supervision: MRi, SA, HR, RT, MC, DEG, DC, BMG, LR; Validation: KG, AM, NW, OCW, CR, DH, MRo, DL, VK and BMG; Visualization: KG, AM and FP; Writing – original draft: MRi, KG, AM, FP, MC, DC,BMG, LR; Writing - review & editing: MRi, KG, AM, SA, OCW, LV, MK, FP, NW, CR, DH, TL, HR, RT, MRo, DL, PK, TB, BF, MM, GD, AD, SM, MC, DEG, DC, BMG, LR

## Data Sharing Statement

Anonymized data that underlie the results reported in this article are available upon justified request to the corresponding author.

## References

1. WHO. WHO. Director-General’s opening remarks at the media briefing on COVID-19. Available: https://www.who.int/dg/speeches/detail/whodirector-general-s-opening-remarks-at-the-media-briefing-on-covid-19---11-march-2020

2. Zhu N, Zhang D, Wang W, Li X, Yang B, Song J, et al. A Novel Coronavirus from Patients with Pneumonia in China, 2019. N Engl J Med. 2020;382: 727–733. doi:10.1056/nejmoa2001017

3. WHO. WHO_weekly-operational-update-on-covid-19 29-march-2021. 2021 [cited 30 Mar 2021] p. https://www.who.int/publications/m/item/weekly-ope. Available: https://www.who.int/publications/m/item/weekly-operational-update-on-covid-19---29-march-2021

4. Lake MA. What we know so far: COVID-19 current clinical knowledge and research. Clin Med J R Coll Physicians London. 2020;20: 124–127. doi:10.7861/clinmed.2019-coron

5. Bhopal R. Covid-19 worldwide: We need precise data by age group and sex urgently. BMJ. 2020;369: 32188598. doi:10.1136/bmj.m1366

6. Health E for public, Sanità IS di. Gender differences in COVID-19: the importance of sex-disaggregated data. Available: https://www.epicentro.iss.it/en/coronavirus/sars-cov-2-gender-differences-importance-sex-disaggregated-data

7. Risch M, Grossmann K, Aeschbacher S, Weideli OC, Kovac M, Pereira F, et al. Investigation of the use of a sensor bracelet for the presymptomatic detection of changes in physiological parameters related to COVID-19: an interim analysis of a prospective cohort study (COVI- GAPP). BMJ Open. 2022;12: e058274. doi:10.1136/bmjopen-2021-058274

8. Peckham H, de Gruijter NM, Raine C, Radziszewska A, Ciurtin C, Wedderburn LR, et al. Male sex identified by global COVID-19 meta-analysis as a risk factor for death and ITU admission. Nat Commun. 2020;11: 1–10. doi:10.1038/s41467-020-19741-6

9. Williamson EJ, Walker AJ, Bhaskaran K, Bacon S, Bates C, Morton CE, et al. Factors associated with COVID-19-related death using OpenSAFELY. Nature. 2020;584: 430–436. doi:10.1038/s41586-020-2521-4

10. Vikse J, Lippi G, Henry BM. Do sex-specific immunobiological factors and differences in angiotensin converting enzyme 2 (ACE2) expression explain increased severity and mortality of COVID-19 in males? Diagnosis (Berlin, Ger. 2020;7: 385–386. doi:10.1515/dx-2020-0054

11. Gebhard C, Regitz-Zagrosek V, Neuhauser HK, Morgan R, Klein SL. Impact of sex and gender on COVID-19 outcomes in Europe. Biol Sex Differ. 2020;11: 29. doi:10.1186/s13293-020-00304-9

12. Aggarwal NR, Patel HN, Mehta LS, Sanghani RM, Lundberg GP, Lewis SJ, et al. Sex Differences in Ischemic Heart Disease: Advances, Obstacles, and Next Steps. Circ Cardiovasc Qual Outcomes. 2018;11: 1–14. doi:10.1161/CIRCOUTCOMES.117.004437

13. Jacobsen H, Klein SL. Sex Differences in Immunity to Viral Infections. Frontiers in Immunology. 2021. p. 3483. Available: https://www.frontiersin.org/article/10.3389/fimmu.2021.720952

14. Oertelt-Prigione S. Immunology and the menstrual cycle. Autoimmun Rev. 2012;11: A486–92. doi:10.1016/j.autrev.2011.11.023

15. Kovacevic V, Markovic A, Veen D, Brakenhoff TB, Mitratza M, Goodale BM. Menstrual Cycle Influence on COVID-19 Detection Using Machine Learning and a Wearable Medical Device [A120]. Obstet Gynecol. 2022;139. Available: https://journals.lww.com/greenjournal/Fulltext/2022/05001/Menstrual_Cycle_Influence_on_COVID_19_Detection.118.aspx

16. Takahashi T, Ellingson MK, Wong P, Israelow B, Lucas C, Klein J, et al. Sex differences in immune responses that underlie COVID-19 disease outcomes. Nature. 2020;588: 315–320. doi:10.1038/s41586-020-2700-3

17. Bogu GK, Snyder MP. Deep learning-based detection of COVID-19 using wearables data. medRxiv. 2021; 2021.01.08.21249474. Available: http://medrxiv.org/content/early/2021/01/09/2021.01.08.21249474.abstract

18. Mishra T, Wang M, Metwally AA, Bogu GK, Brooks AW, Bahmani A, et al. Pre-symptomatic detection of COVID-19 from smartwatch data. Nat Biomed Eng. 2020;4: 1208–1220. doi:10.1038/s41551-020-00640-6

19. Brakenhoff TB, Franks B, Goodale BM, van de Wijgert J, Montes S, Veen D, et al. A prospective, randomized, single-blinded, crossover trial to investigate the effect of a wearable device in addition to a daily symptom diary for the remote early detection of SARS- CoV-2 infections (COVID-RED): a structured summary of a study protocol fo. Trials. 2021;22: 1–5. doi:10.1186/s13063-021-05241-5

20. Quer G, Radin JM, Gadaleta M, Baca-Motes K, Ariniello L, Ramos E, et al. Wearable sensor data and self-reported symptoms for COVID-19 detection. Nat Med. 2021;27: 73–77. doi:10.1038/s41591-020-1123-x

21. Ates HC, Yetisen AK, Güder F, Dincer C. Wearable devices for the detection of COVID-19. Nat Electron. 2021;4: 13–14. doi:10.1038/s41928-020-00533-1

22. Cosoli G, Scalise L, Poli A, Spinsante S. Wearable devices as a valid support for diagnostic excellence: lessons from a pandemic going forward. Health Technol (Berl). 2021;11: 673–675. doi:10.1007/s12553-021-00540-y

23. Risch L, Conen D, Aeschbacher S, Grossmann K RM. Defining the role of a fertility bracelet for early recognition and monitoring of COVID-19 in Liechtenstein: an observational study (COVI-GAPP). In: 10. April 2020. p. 10.1186/ISRCTN51255782.

24. Hamvas G, Hofmann A, Sakalidis V, Goodale B, Shilaih M, Leeners B. Innovative Trial Design Using Digital Approaches: An Example From Reproductive Medicine [21I]. Obstet Gynecol. 2020;135. Available: https://journals.lww.com/greenjournal/Fulltext/2020/05001/Innovative_Trial_Design_Using_Digital_Approaches_.338.aspx

25. Conen D, Schön T, Aeschbacher S, Paré G, Frehner W, Risch M, et al. Genetic and phenotypic determinants of blood pressure and other cardiovascular risk factors: Methodology of a prospective, population-based cohort study. Swiss Med Wkly. 2013;143: 1–9. doi:10.4414/smw.2013.13728

26. Goodale BM, Shilaih M, Falco L, Dammeier F, Hamvas G, Leeners B. Wearable sensors reveal menses-driven changes in physiology and enable prediction of the fertile window: Observational study. J Med Internet Res. 2019;21. doi:10.2196/13404

27. French-Mowat E, Burnett J. How are medical devices regulated in the European Union? J R Soc Med. 2012;105 Suppl: 22–28. doi:10.1258/jrsm.2012.120036

28. Muehlematter UJ, Daniore P, Vokinger KN. Approval of artificial intelligence and machine learning-based medical devices in the USA and Europe (2015–20): a comparative analysis. Lancet Digit Heal. 2021;3: e195–e203. doi:10.1016/S2589-7500(20)30292-2

29. Degenhardt L, Charlson F, Ferrari A, Santomauro D, Erskine H, Mantilla-Herrara A, et al. The global burden of disease attributable to alcohol and drug use in 195 countries and territories, 1990–2016: a systematic analysis for the Global Burden of Disease Study 2016. The Lancet Psychiatry. 2018;5: 987–1012. doi:10.1016/S2215-0366(18)30337-7

30. Risch M, Weber M, Thiel S, Grossmann K, Wohlwend N, Lung T, et al. Temporal course of SARS-CoV-2 antibody positivity in patients with COVID-19 following the first clinical presentation. medRxiv. 2020;2020. doi:10.1101/2020.10.17.20214445

31. R Core Team (2021). The R Project for Statistical Computing. Vienna. Austria: R Foundation for Statistical Computing.; Available: https://www.r-project.org/

32. Van Rossum, G., & Drake FL. Python 3 Reference Manual. Scotts Valley, CA: CreateSpace.; 2009.

33. Shi Y, Wang Y, Shao C, Huang J, Gan J, Huang X, et al. COVID-19 infection: the perspectives on immune responses. Cell Death Differ. 2020;27: 1451–1454. doi:10.1038/s41418-020-0530-3

34. Junqueira C, Crespo Â, Ranjbar S, de Lacerda LB, Lewandrowski M, Ingber J, et al. FcγR-mediated SARS-CoV-2 infection of monocytes activates inflammation. Nature. 2022;606: 576–584. doi:10.1038/s41586-022-04702-4

35. Webster JI, Tonelli L, Sternberg EM. Neuroendocrine regulation of immunity. Annu Rev Immunol. 2002;20: 125–163. doi:10.1146/annurev.immunol.20.082401.104914

36. Ernst G. Heart-Rate Variability-More than Heart Beats? Front public Heal. 2017;5: 240. doi:10.3389/fpubh.2017.00240

37. Papaioannou V, Pneumatikos I, Maglaveras N. Association of heart rate variability and inflammatory response in patients with cardiovascular diseases: current strengths and limitations. Front Physiol. 2013;4: 174. doi:10.3389/fphys.2013.00174

38. Natarajan A, Su HW, Heneghan C, Blunt L, O’Connor C, Niehaus L. Measurement of respiratory rate using wearable devices and applications to COVID-19 detection. npj Digit Med. 2021;4. doi:10.1038/s41746-021-00493-6

39. Robert P Hirten, MD, Matteo Danieletto, PhD, […], and Zahi A, Fayad P. Use of Physiological Data From a Wearable Device to Identify SARS-CoV-2 Infection and Symptoms and Predict COVID-19 Diagnosis: Observational Study. J Med Internet Res. 2021; https://www.ncbi.nlm.nih.gov/pmc/articles/PMC79015.

40. Conroy B, Silva I, Mehraei G, Damiano R, Gross B, Salvati E, et al. Real-time infection prediction with wearable physiological monitoring and AI to aid military workforce readiness during COVID-19. Sci Rep. 2022;12: 3797. doi:10.1038/s41598-022-07764-6

41. Marianna Mitratza, MD, PhD Ms, Brianna Mae Goodale P, Aizhan Shagadatova Ms, Vladimir Kovacevic P, Janneke van de Wijgert, MD, PhD M, Timo B. Brakenhoff P, et al. The Performance of Wearable Sensors in the Detection of SARS-CoV-2 infection: A Systematic Review. Lancet Digit Heal. 2022.

42. Shaw LJ, Pepine CJ, Xie J, Mehta PK, Morris AA, Dickert NW, et al. Quality and Equitable Health Care Gaps for Women: Attributions to Sex Differences in Cardiovascular Medicine. J Am Coll Cardiol. 2017;70: 373–388. doi:10.1016/j.jacc.2017.05.051

43. Schumacher Dimech A, Ferretti MT, Sandset EC, Santuccione Chadha A. The role of sex and gender differences in precision medicine: the work of the Women’s Brain Project. Eur Heart J. 2021;42: 3215–3217. doi:10.1093/eurheartj/ehab297

44. Yu AYX, Hill MD, Asdaghi N, Boulanger J-M, Camden M-C, Campbell BC V, et al. Sex Differences in Diagnosis and Diagnostic Revision of Suspected Minor Cerebral Ischemic Events. Neurology. 2021;96: e732–e739. doi:10.1212/WNL.0000000000011212

45. Cirillo D, Catuara-Solarz S, Morey C, Guney E, Subirats L, Mellino S, et al. Sex and gender differences and biases in artificial intelligence for biomedicine and healthcare. npj Digit Med. 2020;3: 81. doi:10.1038/s41746-020-0288-5

